# Most Hospitalized Patients with Significant Tricuspid Regurgitation Have Advanced Disease Preventing Transcatheter Tricuspid Valve Intervention

**DOI:** 10.1101/2023.09.14.23295588

**Authors:** Allison O. Dumitriu Carcoana, Christopher B. Scoma, Sebastian N. Maletz, Jose A. Malavet, Daniela R. Crousillat, Fadi A. Matar

## Abstract

**Background:** More than moderate tricuspid regurgitation (TR) is associated with high mortality. Surgical tricuspid valve repair and replacements are rarely performed due to high operative mortality risk, mainly attributed to late presentation. Novel transcatheter tricuspid valve intervention (TTVI) devices are being developed as an alternative to surgery. The population of patients presenting to tertiary care centers who can benefit from TTVI has not been well defined. Our objective was to study the characteristics of these patients and to define the sub-population who may be candidates for TTVI.

**Methods:** We analyzed 12,677 consecutive 2D echocardiograms completed at our tertiary care center between March 2021 and March 2022 and identified 581 inpatients with more than moderate TR. Clinical and echocardiographic data were collected by individual chart review. The 2021 European Society of Cardiology (ESC) guidelines on the management of valvular disease are currently the only guidelines that include TTVI. We used these guidelines to retrospectively assign the 581 patients to medical therapy, surgery, or TTVI treatment.

**Results:** 470 patients (80.9%) were assigned to medical therapy, 57 (9.8%) were assigned to TTVI, and 54 (9.3%) were assigned to tricuspid valve surgery. Of note, 76.2% (443/581) of patients were precluded from any intervention due to advanced disease or anatomic ineligibility, and only 4.6% (27/581) presented too early for intervention, being both asymptomatic and without RV dilatation.

**Conclusion:** Only 9.8 percent of patients presenting to a tertiary care center with significant TR would be hypothetical candidates for TTVI when these technologies are approved in the United States. Earlier identification and treatment of TR could increase the number of patients who may benefit from interventions including TTVI.

**CLINICAL PERSPECTIVE:** *What is New?:*

- Most patients presenting to a tertiary care center with significant TR are too advanced for any intervention and have high in-hospital and late mortality rates.
- Tertiary care centers planning to offer TTVI should expect that, based on the current ESC guidelines and late presentation of TR, approximately 10% of patients with more than moderate TR will be TTVI candidates.

*What are the clinical implications?:*

- There is a need to improve care for patients with TR through earlier detection and referral for intervention.
- Future clinical trials with longer follow up will be needed to assess TTVI devices in patients at earlier or even asymptomatic stages of TR when there may be a greater likelihood of improving survival.

## INTRODUCTION

More than moderate tricuspid regurgitation (TR) is associated with high operative mortality.^1–8^ Due to significant operative risk, surgical TVR is uncommonly performed due in part to late clinical presentation.^9–12^ Several new transcatheter tricuspid valve intervention (TTVI) devices are being developed as a lower-risk alternative to surgery. Many of these devices are in use in Europe and are in the process of being evaluated for approval in the United States.^13^ The prevalence of moderate and severe TR in the United States is estimated at 1,600,000.^14^ However, the population of TR patients who can benefit from TTVI is yet to be defined.

Furthermore, The European Society of Cardiology (ESC) 2021 guidelines for the management of valvular heart disease are the only guidelines that currently include TTVI as a treatment option.^15^ As these therapies become available in the United States^13^, it is essential to understand the patient populations who may best benefit. In this study, we examined the clinical and echocardiographic variables of patients with significant TR presenting to a tertiary care center and defined the sub-population who may be candidates for TTVI, based on the ESC guidelines.

## METHODS

We retrospectively identified 12,677 consecutive patients age ≥18 years who had a transthoracic echocardiogram performed at our center between March 2021 and March 2022. Of these, we collected the clinical and demographic information of 581 patients with more than moderate TR. A consistent group of cardiologists graded TR severity. To further justify our analysis, two cardiologists (C.B.S., D.R.C.) independently re-reviewed 10% of the echocardiograms. The difference of proportions was calculated to validate reliability of categorical parameters prone to some degree of subjectivity. The potentially subjective parameters of importance to our study were the presence of more than moderate TR and the presence of severe right ventricular (RV) dysfunction. For TR severity, the difference of proportions was 0.05 with a standard error of 0.03 and a p-value of 0.0793. For RV dysfunction, the standard error was 0.12 ± 0.07 with a p-value of 0.0791. Thus, we cannot conclude that there was a significant difference between the original and repeated echocardiogram reviews, substantiating consistent grading of these parameters.

We used the process described below to place patients into one of three retrospectively assigned treatment groups: medical therapy, surgery, or TTVI (Figure 1).^15^ The retrospectively assigned treatments were not necessarily those applied in actual practice. Our Institutional Review Board (USF IRB Administrative Review; FWA No. 00001669) determined that this protocol met the criteria for exemption from IRB review. One author (F.A.M.) had full access to all the data in the study and takes responsibility for its integrity and the data analysis.

**Figure 1.**
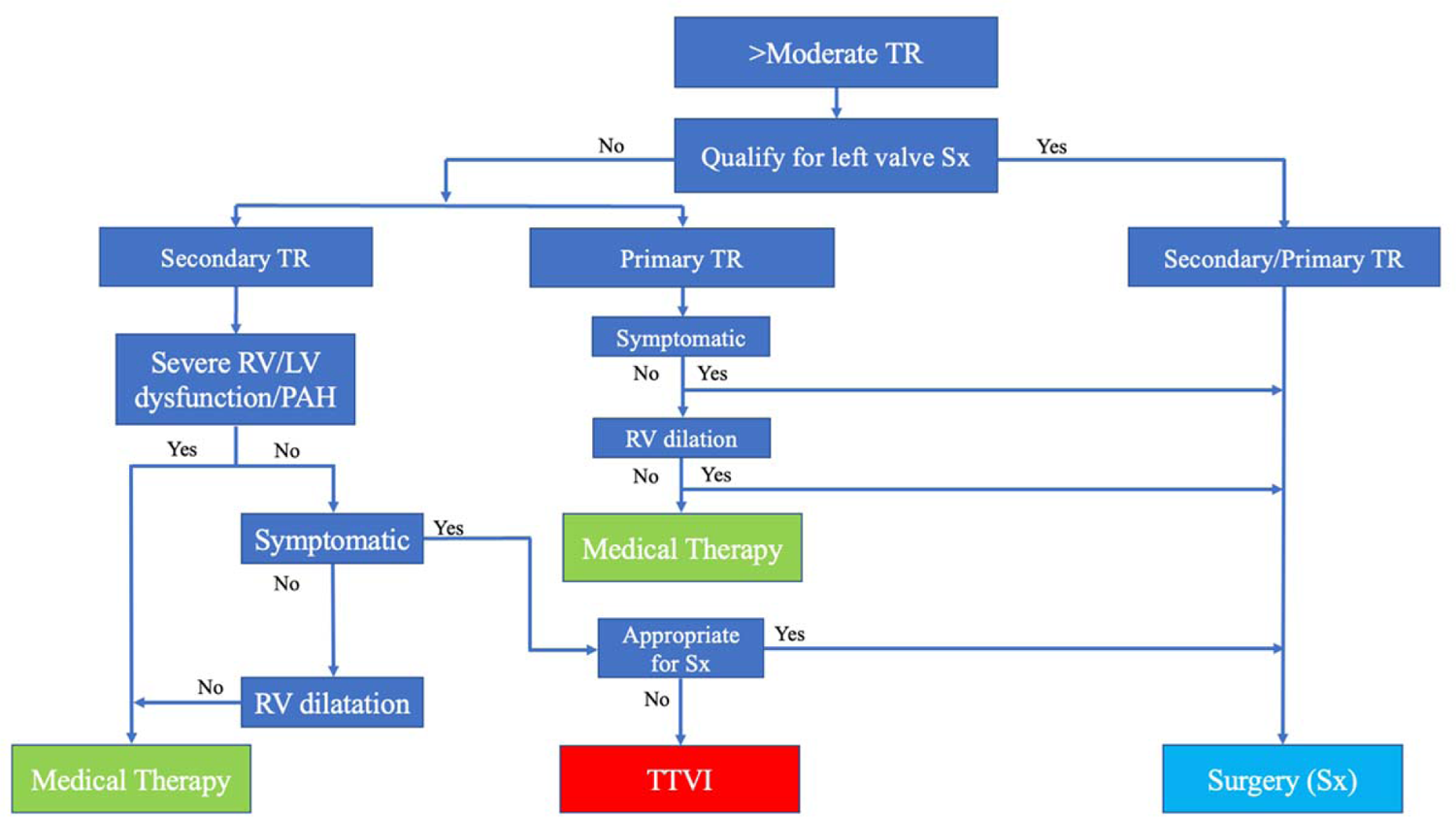
Management of tricuspid regurgitation based on the 2021 ESC guidelines The 2021 European Society of Cardiology (ESC) guidelines for the management of valvular disease include an algorithm for determining the management of tricuspid regurgitation (TR): medical therapy, transcatheter tricuspid valve intervention (TTVI) or surgery (Sx). RV dilatation = right ventricular dilatation; Severe RV/LV dysfunction/PAH= severe right ventricular or left ventricular dysfunction or severe pulmonary arterial hypertension.

### Definitions

#### Modified ESC Algorithm

The ESC guidelines recommend that a multidisciplinary Heart Team should determine patients’ candidacy for surgery vs. TTVI. In clinical practice, all patients under consideration for TV surgery would be evaluated based on their clinical characteristics. However, the ESC guidelines do not specify the criteria that should be used for surgical candidacy. Our hypothetical Heart Team identified patients with age ≥80 or prior history of open-heart surgery, valve surgery, cirrhosis, or end-stage renal disease as too high risk for surgery. Patients considered for surgery based on ESC criteria who were too high risk based on Heart Team criteria were assigned to TTVI. Patients with primary TR who were too high-risk for surgery and not anatomically viable candidates for TTVI based on endocarditis or congenital TR etiology were assigned to medical therapy. Our modifications to the ESC algorithm are depicted in Figure 2 and Figure 3.

**Figure 2.**
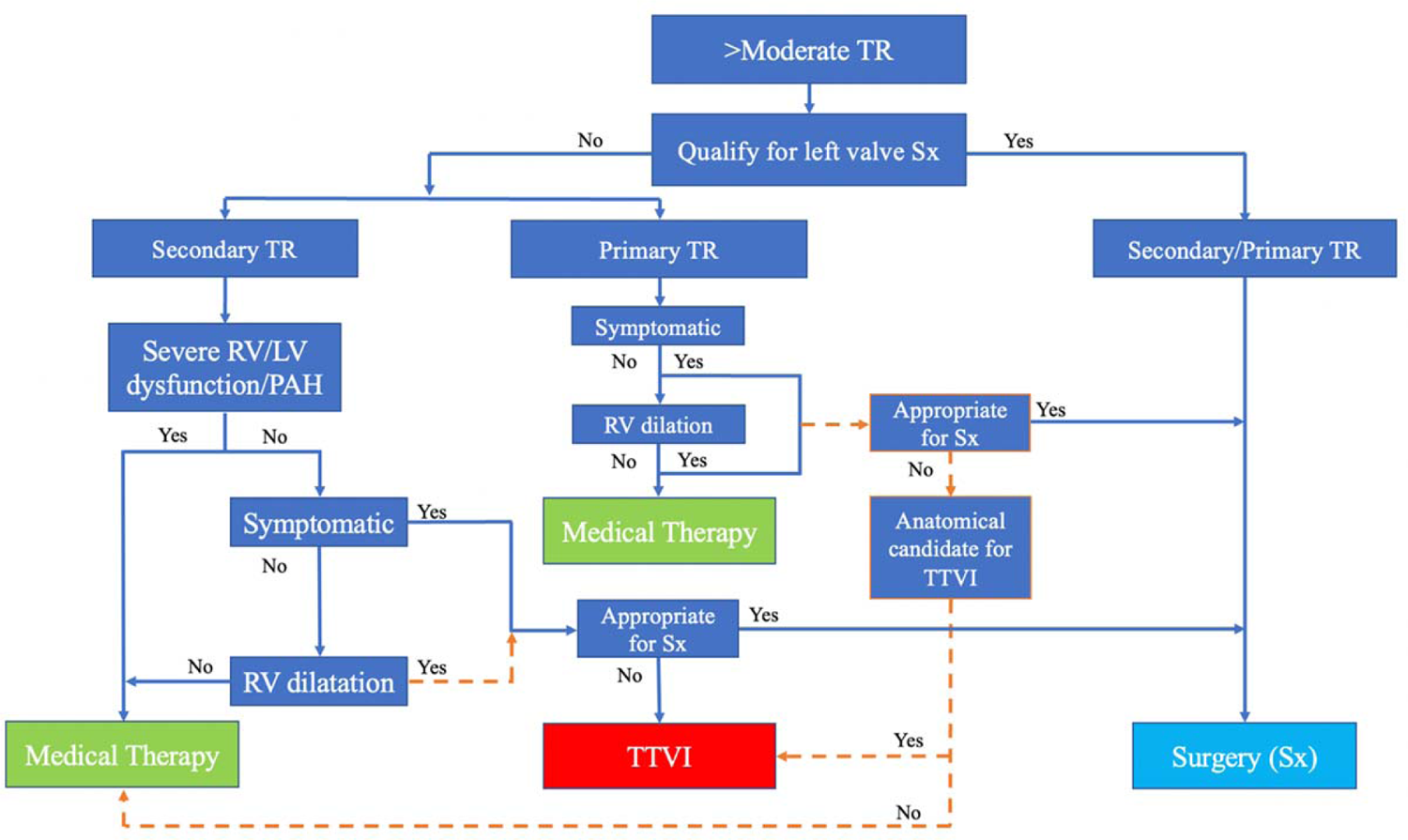
The modified ESC algorithm We modified the 2021 ESC algorithm for determining the management of tricuspid regurgitation (orange dashed lines and orange borders). Abbreviations as in Figure 1.

**Figure 3.**
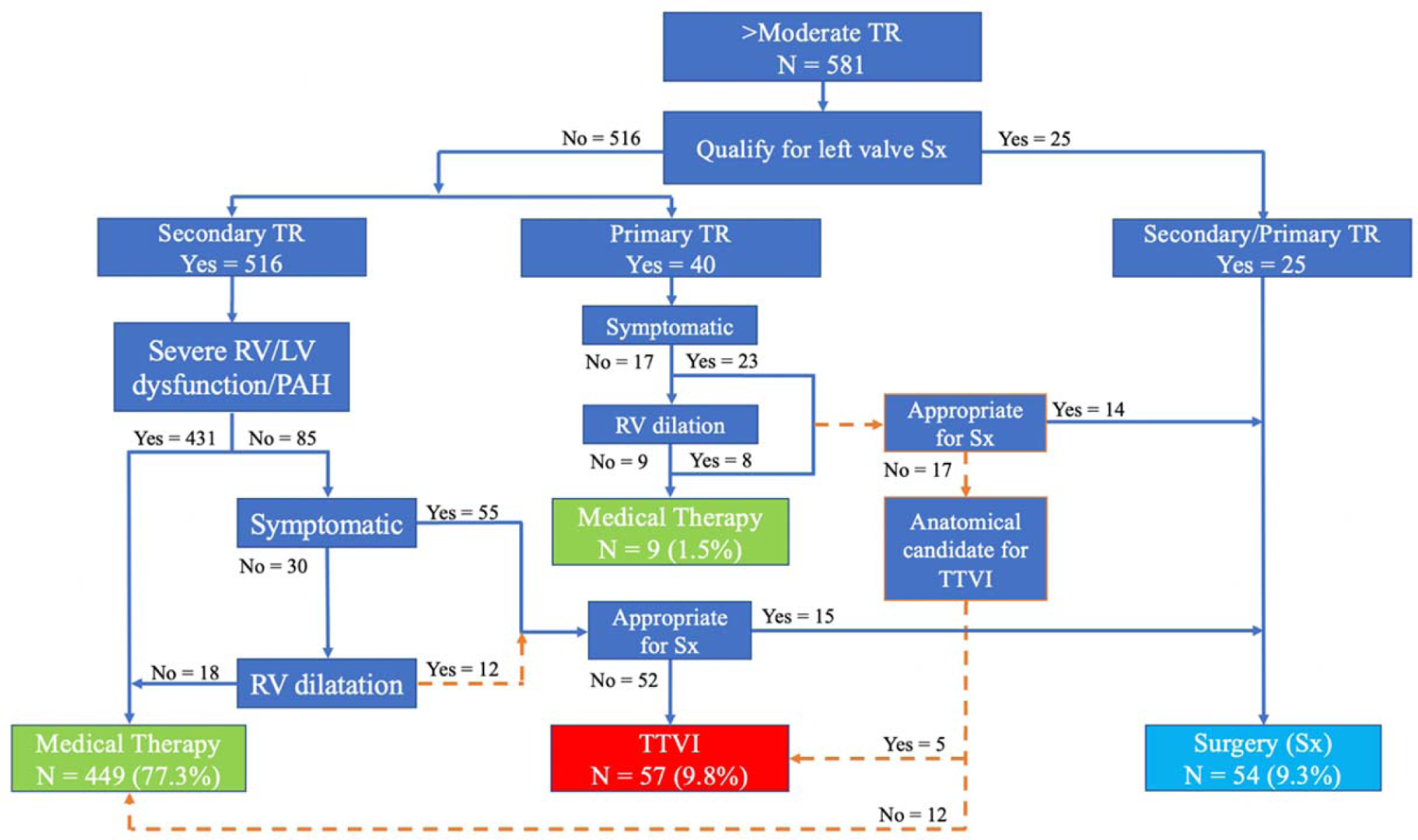
Applying the modified ESC algorithm We applied the modified European Society of Cardiology (ESC) algorithm to our cohort of 581 patients with more than moderate TR and determined the numbers of patients who were candidates for medical therapy, TTVI, or Sx. Abbreviations as in Figure 1.

#### Need for Left-Sided Valve Surgery

The first step was to determine whether patients needed left-sided valve surgery, criteria not well-defined in the ESC guidelines. We defined qualification for left-sided valve surgery as >moderate mitral or aortic valve regurgitation or stenosis plus meeting Heart Team criteria. The patients who did not qualify were then divided by primary and secondary TR etiologies.

#### Primary TR Without Left Sided Valve Dysfunction

Primary TR included endocarditis, carcinoid, lead impingement, and congenital causes. The ESC guidelines recommend first determining whether these patients had symptoms of right heart failure (RHF). NYHA class was not available for over half of our cohort, therefore; we defined signs of RHF as lower extremity edema or ascites around the time of the echocardiogram. Asymptomatic patients were then assessed for RV dilatation. RV dilatation was determined in accordance with the 2015 American Society of Echocardiography (ASE) recommendations for cardiac chamber quantification, including basal diameter >41mm, mid diameter >35mm, or longitudinal diameter >83mm.^16^ Asymptomatic patients without RV dilatation were assigned to medical therapy. Patients with symptoms or RV dilatation were evaluated for surgical candidacy. Those who qualified were assigned to tricuspid valve surgery, while those at too high a surgical risk were assigned to TTVI.

#### Secondary TR Without Left Sided Valve Dysfunction

The ESC guidelines recommend that for the patients who do not qualify for left-sided surgery and have secondary TR: severe RV dysfunction, severe LV dysfunction, and severe pulmonary hypertension are exclusion criteria from any intervention, surgical or transcatheter. We assessed the degree of dysfunction based on the echocardiograms using criteria from the 2015 ASE recommendations.^16^ Severe RV dysfunction was qualitatively described and confirmed quantitively with a TAPSE <17 mm or an S’ <10 cm/s. Severe LV dysfunction was defined as LVEF <30%. Severe pulmonary hypertension was defined as right ventricular systolic pressure >55 mmHg or TR jet velocity >3.6m/s with systolic septal flattening. The patients who met at least one criterion had advanced disease preventing the use of any intervention and were assigned to the medical therapy group.

Of the remaining patients, those who were asymptomatic with no RV dilatation were assigned to medical therapy. Those who were symptomatic or asymptomatic with RV dilatation were assigned to either surgery or TTVI based on Heart Team criteria.

#### Early vs. Late Presentation

We divided the medical therapy group by early and late presenters, so that TR etiology and mortality data could be more accurately assessed with respect to disease severity. Patients without need for left-sided valve surgery who have secondary TR without severe LV dysfunction, severe RV dysfunction, severe pulmonary hypertension, symptoms of RHF, and RV dilatation were assigned to medical therapy. Similarly, patients with primary TR without symptoms of RHF and without RV dilatation were assigned to medical therapy. These two groups were therefore assigned to medical therapy due to early presentation.

On the other hand, patients with severe LV or RV dysfunction or severe pulmonary hypertension were precluded from any intervention and were therefore assigned to medical therapy due to advanced stage disease. In our modified ESC algorithm, we also considered the primary TR patients assigned to medical therapy due to high surgical risk and anatomical unviability for TTVI as advanced.

### TR Etiology

We determined the subtype of functional TR based on the process described by Topilsky et al.^17^ Left-sided valvular disease was defined as greater than moderate mitral or aortic valve regurgitation or stenosis. LV systolic dysfunction was defined as LVEF <40% and the absence of moderate-severe left valvular disease. Severe pulmonary hypertension was defined as pulmonary arterial pressure >55 mmHg or TR jet velocity >3.6m/s with systolic septal flattening and the absence of both moderate-severe left sided valvular disease and LV systolic dysfunction. Isolated TR was defined by the absence of severe pulmonary hypertension, moderate-severe left valvular disease, and LV systolic dysfunction.

### Statistics

The standard error of proportions was calculated between the original and repeated assessments of TR severity and RV dysfunction for 10% of the echocardiograms included in this study. This was done to validate the reliability of these parameters because they may be prone to some degree of subjectivity. We reported mean ± standard deviation or median and first and third quartile (Q1, Q3) values for continuous variables and frequency (percentage) for categorical variables. Variables that did not follow a normal distribution curve were reported as median (Q1, Q3). Cox regression analysis was used for survival analysis, presented as the median survival time with standard deviation and 95% confidence intervals. Kaplan-Meier curves were generated to compare overall survival between the whole cohort compared to survival curves of each treatment group, with medical therapy divided into early and advanced groups. IBM SPSS Statistics for Macintosh, Version 29.0, was used for survival analysis.

## RESULTS

### Patient Characteristics

Table 1 displays a summary of all clinical data collected for the 581 patients with more than moderate TR. In this cohort, 52.3% (304/581) were women, 56.3% (325/577) were White, and the average age was 65.7 years. Of note, lower extremity edema was present in 67.8% (394/581), ascites in 34.3% (199/581), cirrhosis in 17.7% (103/581), and atrial fibrillation or flutter in 60.9% (354/581). Of the patients who had NYHA class noted in their chart, 78.5% (234/298) were NYHA class III or IV. Table 2 displays the echocardiographic parameters of the 581 patients. 92.8% (539/581) had functional TR. The median TAPSE was 15 mm, median RVSP 58 mmHg, and 57.0% (306/537) had severe pulmonary hypertension.

**Table 1.**
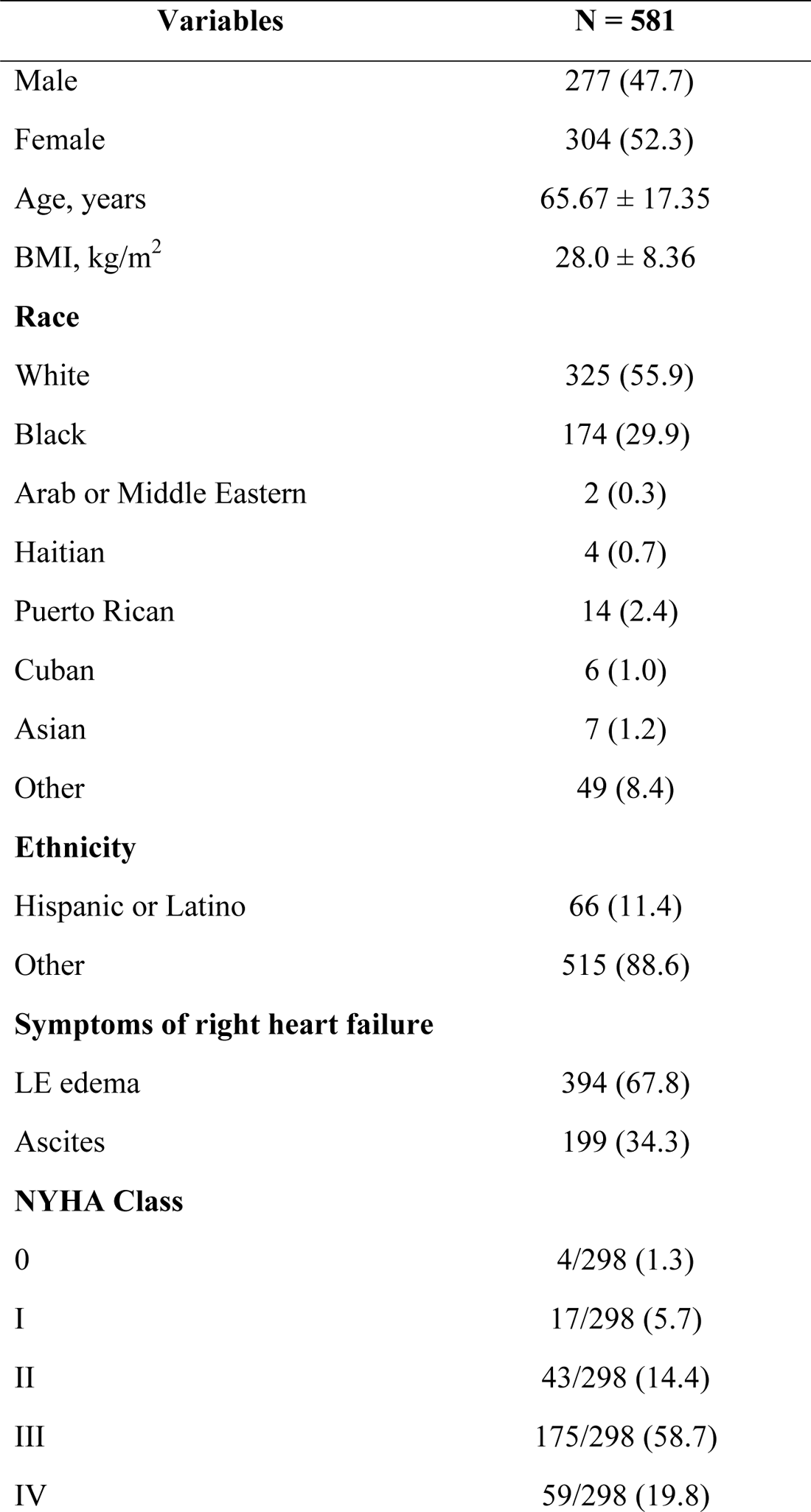
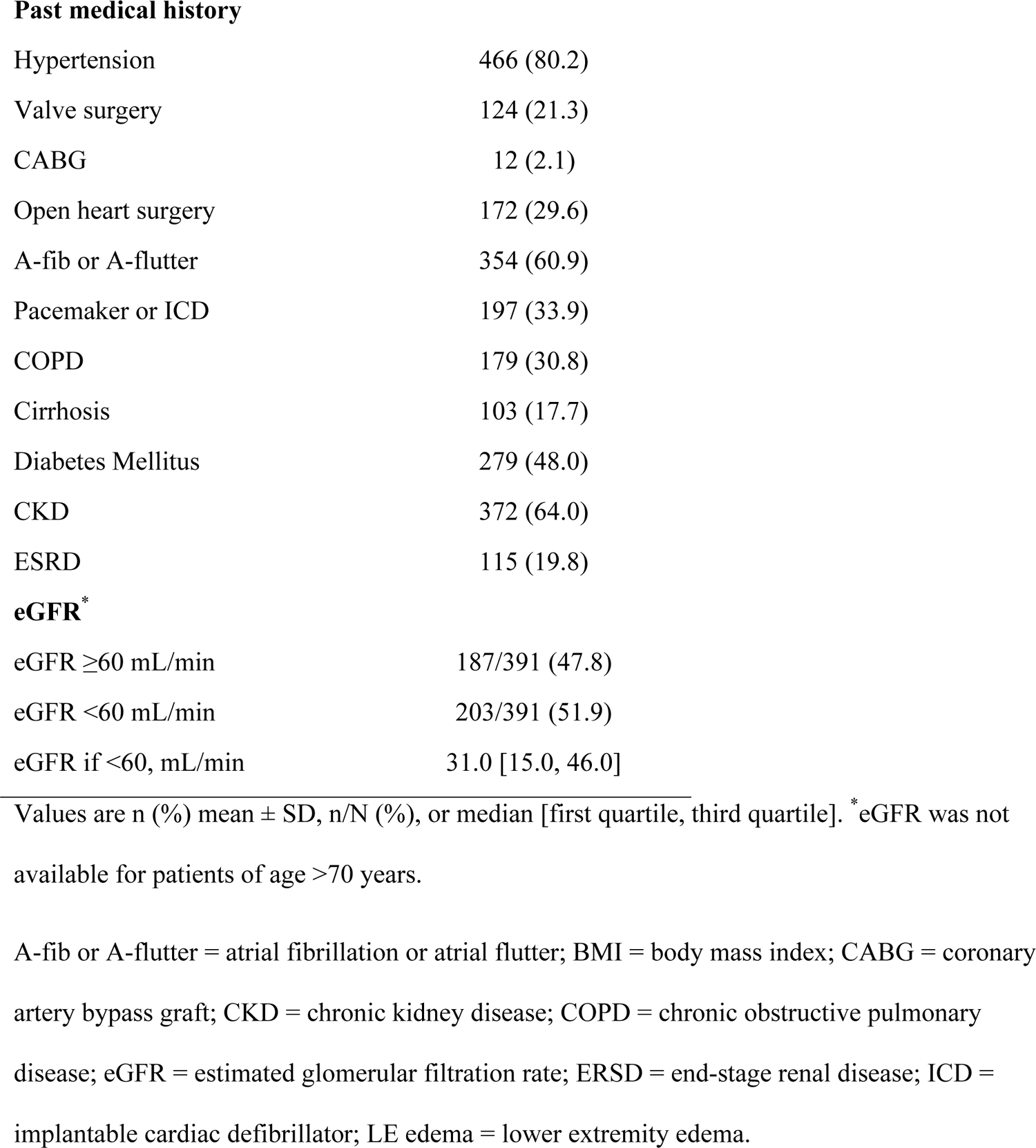
Baseline clinical characteristics of the total cohort.

**Table 2.**
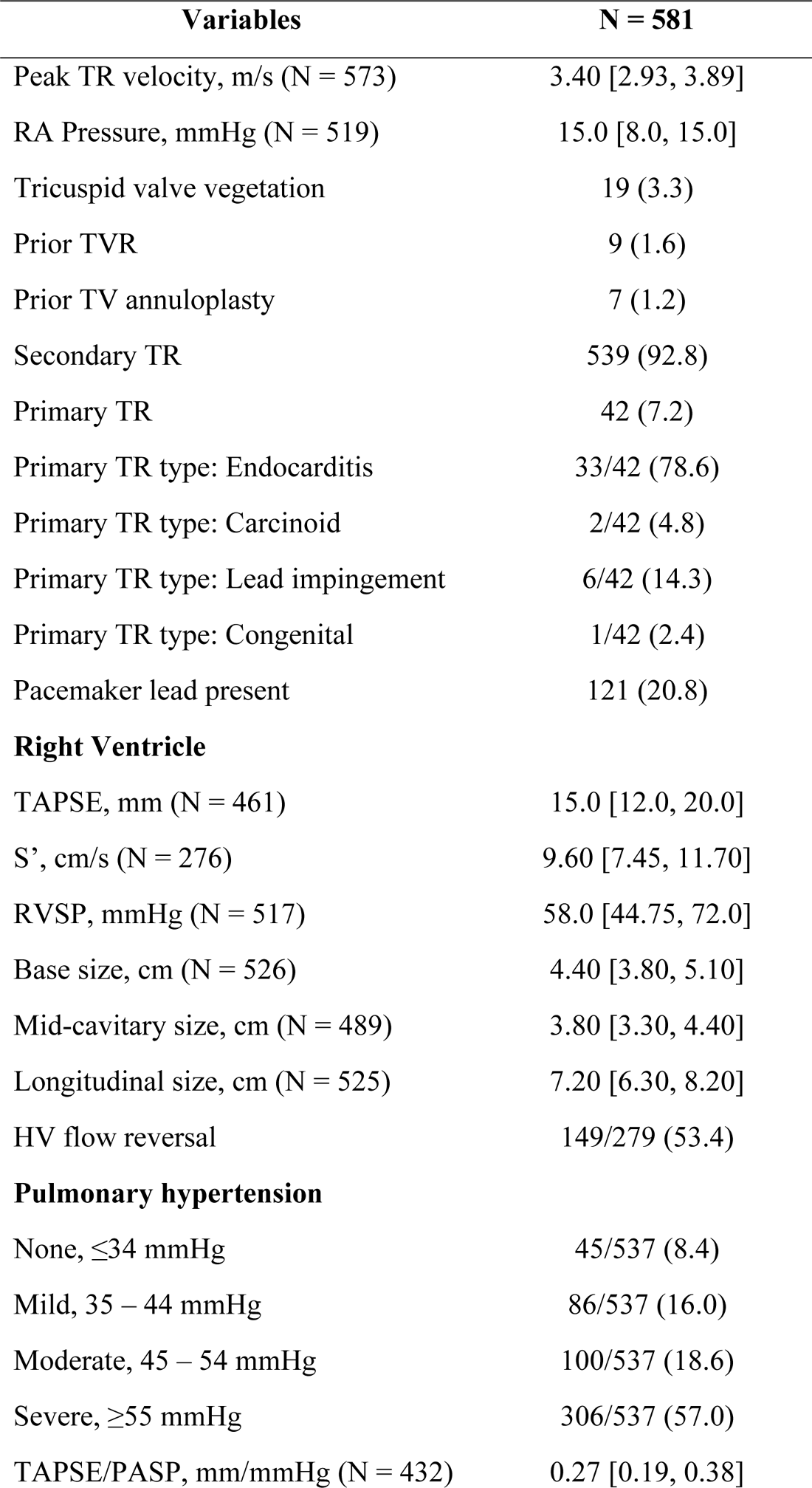
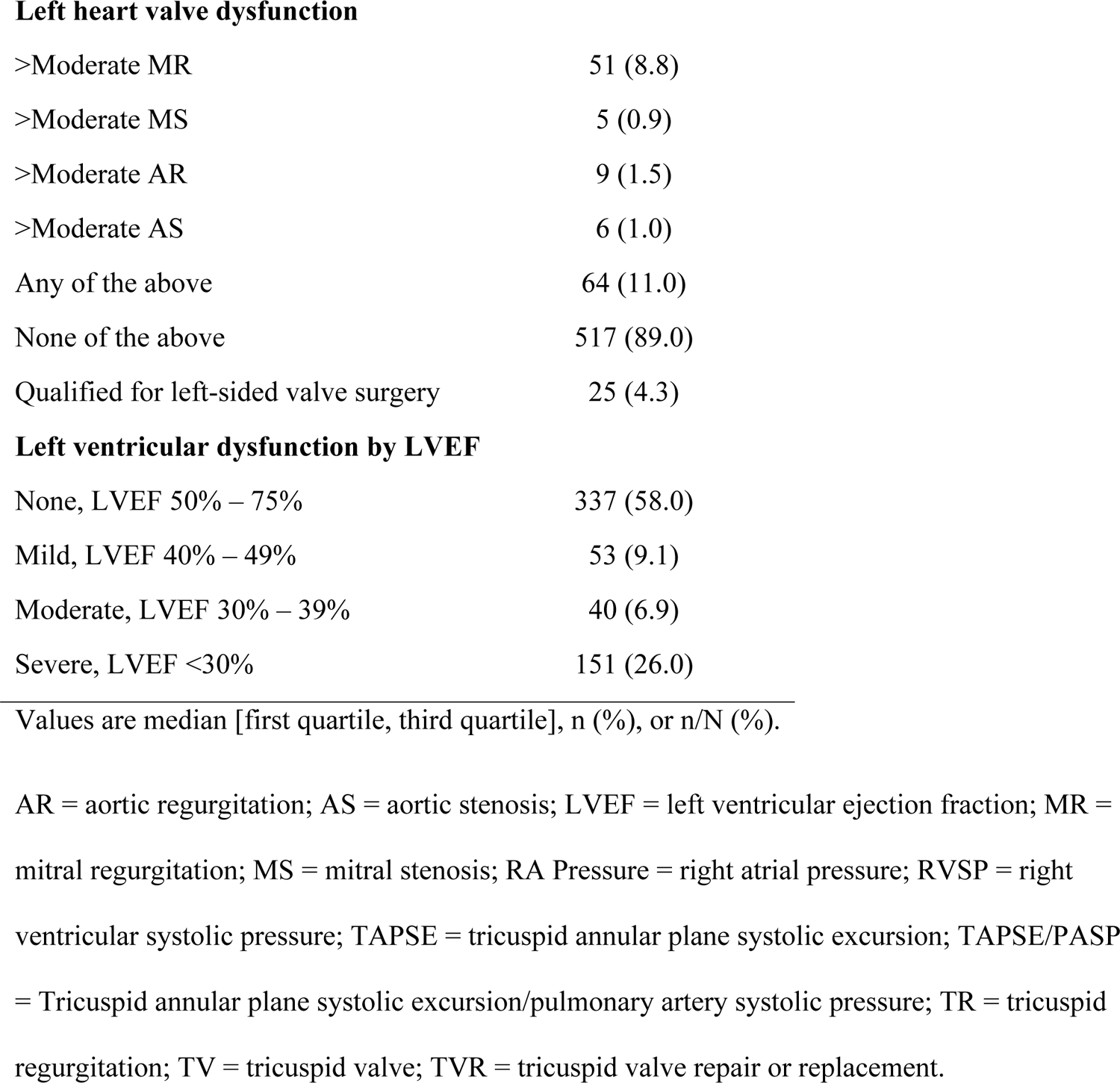
Echocardiographic parameters of the total cohort.

| **Variables** | **N = 581** |
| --- | --- |
| Peak TR velocity, m/s (N = 573) | 3.40 [2.93, 3.89] |
| RA Pressure, mmHg (N = 519) | 15.0 [8.0, 15.0] |
| Tricuspid valve vegetation | 19 (3.3) |
| Prior TVR | 9 (1.6) |
| Prior TV annuloplasty | 7 (1.2) |
| Secondary TR | 539 (92.8) |
| Primary TR | 42 (7.2) |
| Primary TR type: Endocarditis | 33/42 (78.6) |
| Primary TR type: Carcinoid | 2/42 (4.8) |
| Primary TR type: Lead impingement | 6/42 (14.3) |
| Primary TR type: Congenital | 1/42 (2.4) |
| Pacemaker lead present | 121 (20.8) |
| **Right Ventricle** |  |
| TAPSE, mm (N = 461) | 15.0 [12.0, 20.0] |
| S', cm/s (N = 276) | 9.60 [7.45, 11.70] |
| RVSP, mmHg (N = 517) | 58.0 [44.75, 72.0] |
| Base size, cm (N = 526) | 4.40 [3.80, 5.10] |
| Mid-cavitary size, cm (N = 489) | 3.80 [3.30, 4.40] |
| Longitudinal size, cm (N = 525) | 7.20 [6.30, 8.20] |
| HV flow reversal | 149/279 (53.4) |
| **Pulmonary hypertension** |  |
| None, $\leq 34$ mmHg | 45/537 (8.4) |
| Mild, 35 – 44 mmHg | 86/537 (16.0) |
| Moderate, 45 – 54 mmHg | 100/537 (18.6) |
| Severe, $\geq 55$ mmHg | 306/537 (57.0) |
| TAPSE/PASP, mm/mmHg (N = 432) | 0.27 [0.19, 0.38] |
| **Left heart valve dysfunction** |  |
| >Moderate MR | 51 (8.8) |
| >Moderate MS | 5 (0.9) |
| >Moderate AR | 9 (1.5) |
| >Moderate AS | 6 (1.0) |
| Any of the above | 64 (11.0) |
| None of the above | 517 (89.0) |
| Qualified for left-sided valve surgery | 25 (4.3) |
| **Left ventricular dysfunction by LVEF** |  |
| None, LVEF 50% – 75% | 337 (58.0) |
| Mild, LVEF 40% – 49% | 53 (9.1) |
| Moderate, LVEF 30% – 39% | 40 (6.9) |
| Severe, LVEF <30% | 151 (26.0) |

Values are median [first quartile, third quartile], n (%), or n/N (%).
AR = aortic regurgitation; AS = aortic stenosis; LVEF = left ventricular ejection fraction; MR = mitral regurgitation; MS = mitral stenosis; RA Pressure = right atrial pressure; RVSP = right ventricular systolic pressure; TAPSE = tricuspid annular plane systolic excursion; TAPSE/PASP = Tricuspid annular plane systolic excursion/pulmonary artery systolic pressure; TR = tricuspid regurgitation; TV = tricuspid valve; TVR = tricuspid valve repair or replacement.

### Treatment Groups Assigned Using the ESC 2021 Guidelines

Figure 3 displays the numbers of patients who were assigned to each treatment group of the modified ESC algorithm.

#### Need for Left-Sided Valve Surgery

64 patients had >moderate mitral regurgitation, mitral stenosis, aortic regurgitation, or aortic stenosis. Of these, 25 qualified for surgery and 39 were too high risk.

#### Primary TR Without Left Sided Valve Dysfunction

40 of the 556 patients who did not qualify for left-sided valve surgery had primary TR. Of these, 23 had symptoms of RHF, and 17 did not. Of the 23 symptomatic patients, 12 were too high risk for surgery. Of these 12, seven had endocarditis (n = 6) or congenital (n = 1) TR etiologies, characterizing anatomic ineligibility for percutaneous treatment via TTVI, and were therefore assigned to medical therapy. The remaining five were appropriate for TTVI with carcinoid (n = 4) or lead impingement (n = 1) TR etiologies.

The 17 asymptomatic patients were assessed for RV dilatation. Eight had RV dilatation, and of these, three were appropriate for surgery. The other five had endocarditis, so they were assigned to medical therapy. The nine remaining patients who were both asymptomatic and without RV dilatation were assigned to medical therapy based on the ESC guidelines.

#### Secondary TR Without Left Sided Valve Dysfunction

The remaining 516 patients who did not qualify for left-sided valve surgery had secondary (functional) TR. Of these, 431 had severe LV or RV dysfunction or severe pulmonary hypertension and were assigned to medical therapy. Of the 85 patients without severe dysfunction, 55 patients were symptomatic; 12 were appropriate for surgery, and 43 were assigned to TTVI instead. Regarding the 30 asymptomatic patients, 18 had no RV dilatation and were assigned to medical therapy. The remaining 12 underwent assessment by Heart Team criteria, and three were ultimately appropriate for surgery, while nine were assigned to TTVI.

#### Assigned Treatment Groups

In total, 470 patients (80.9%) were assigned to medical therapy, 57 (9.8%) were assigned to TTVI, and 54 (9.3%) were assigned to tricuspid valve surgery. TV surgery can be further divided into concomitant (n = 25/54, 46.3.0%) and isolated tricuspid valve surgeries (n = 29/54, 53.7%). Of note, 76.2% (443/581) of patients were precluded from any intervention due to advanced ventricular dysfunction, severe pulmonary hypertension, or anatomic ineligibility, and only 4.6% (27/581) presented too early for intervention, being both asymptomatic and without RV dilatation.

Table S1 describes the clinical characteristics of each assigned treatment group, and Table S2 presents their echocardiographic parameters. Table 3 displays the TR etiologies in each treatment group. In Table 3, we divide the medical therapy group by early and late presentation.

**Table 3.** TR Etiology in the total cohort and by assigned treatment group.

| Parameters | Total cohort | Medical therapy |  | Surgery | TTVI |
| --- | --- | --- | --- | --- | --- |
|  |  | Advanced stage | Early stage |  |  |
|  | N = 581 | N = 443 | N = 27 | N = 54 | N = 57 |
| Primary TR | 42 (7.2) | 12 (2.7) | 9 (33.3) | 16 (29.6) | 5 (8.8) |
| **Functional TR** | 539 (92.8) | 431 (97.3) | 18 (66.7) | 38 (70.4) | 52 (91.2) |
| Left valvular disease | 60 (10.3) | 33 (7.4) | 2 (7.4) | 23 (41.8) | 2 (3.5) |
| LV systolic dysfunction | 157 (27.0) | 150 (33.9) | 1 (3.7) | 1 (1.8) | 5 (8.8) |
| Severe pulmonary HTN | 197 (33.9) | 197 (44.5) | 0 | 0 | 0 |
| Isolated TR | 125 (21.5) | 51 (11.5) | 15 (55.6) | 14 (25.5) | 45 (61.6) |

Values are n (%). Left valvular disease was defined as greater than moderate mitral or aortic valve regurgitation or stenosis. LV systolic dysfunction was defined as LV ejection fraction <40% and absence of moderate-severe left valvular disease. Severe pulmonary hypertension was defined as pulmonary arterial pressure > 55 mmHg or TR jet velocity >3.6m/s with systolic septal flattening and absence of both moderate-severe left valvular disease and LV systolic dysfunction. Isolated TR was defined by the absence of severe pulmonary hypertension, moderate-severe left valvular disease, and LV systolic dysfunction.
HTN = hypertension; LV = left ventricular; TR = tricuspid regurgitation.

### Concomitant CABG

In clinical practice, TV surgery is also performed with concomitant Coronary Artery Bypass Graft surgery (CABG), but it is not included in the 2021 ESC guidelines. In our cohort, 2.9% (17/581) of patients were possible candidates for CABG based on coronary angiography performed within six months of the index echocardiogram. Of these 17 patients, only three remained surgical candidates after applying the Heart Team criteria. Two of these three were already assigned to TV surgery, and the remaining patient had been assigned to medical therapy due to advanced stage. Thus, the addition of concomitant CABG as a consideration for assigning treatment groups in our cohort would result in approximately equal cohort numbers. One patient in our cohort (whom we assigned to surgery) underwent surgical TVR with concomitant CABG during the follow-up period.

### Survival

Each patient in the study had a follow-up time of at least six months or until death. In-hospital and 18-month mortality were highest amongst the patients assigned to TTVI (19.3%, 28.1%) and medical therapy with late presentation (18.5%, 28.9%; Table 4).

**Table 4.**
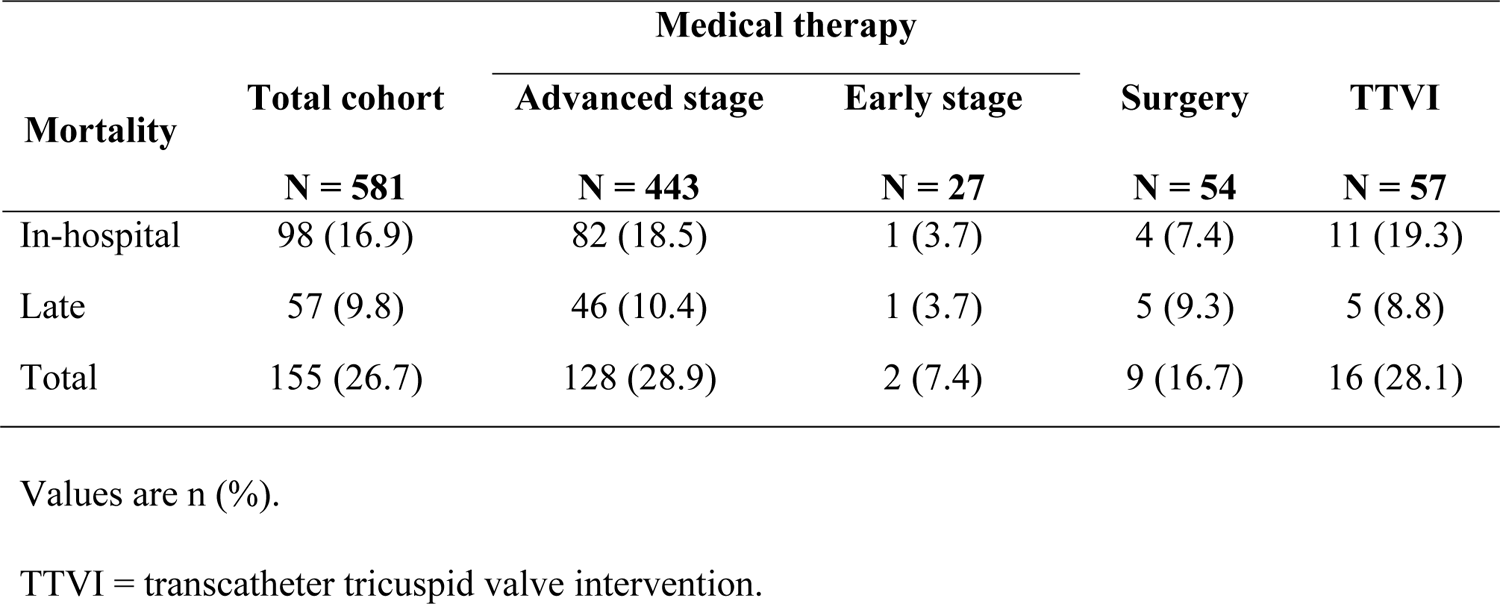
In-hospital and late mortality by treatment group.

In Figure 4, we present the survival curves for the whole cohort compared to survival curves of each treatment group, with medical therapy divided into early and advanced groups. The mean overall survival time in the total cohort was 14.5 months ± a standard error of 0.4 months (95% confidence interval [CI] = 13.74 – 15.31 months). Survival time was shortest in those assigned to TTVI (13.3 ± 1.2 months, 95% CI = 11.0 – 15.6) and medical therapy with advanced stage (14.0 ± 0.5 months, 95% CI = 13.1 – 14.9). Those assigned to medical therapy with early presentation had the longest overall survival (18.1 ± 1.0 months, 95% CI = 16.1 – 20.0), followed by those assigned to surgery (15.1 ± 1,0 months, 95% CI = 13.2 – 17.0).

**Figure 4.**
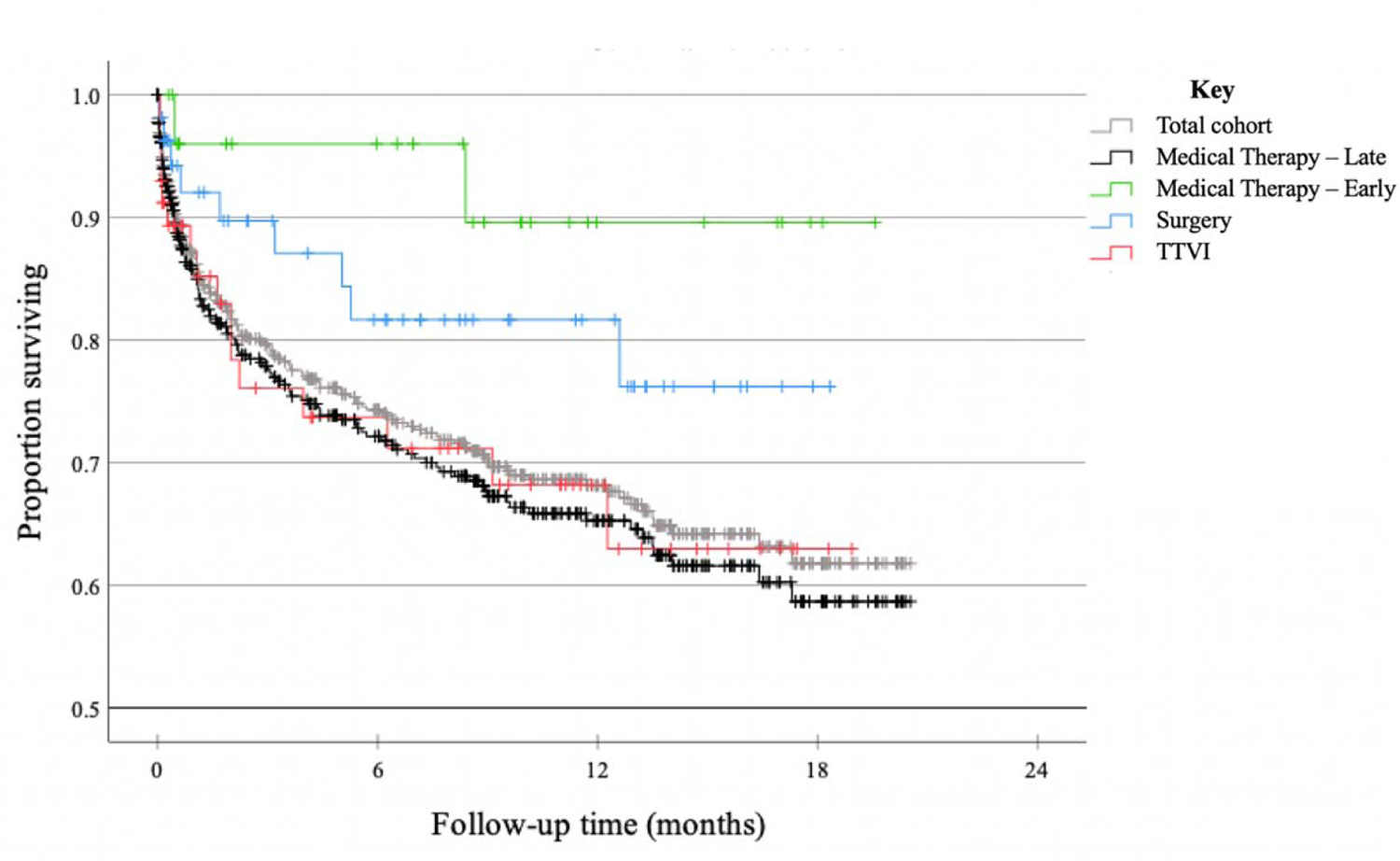
Overall survival in the total cohort and by assigned treatment group. Kaplan-Meier curves of overall survival in the total cohort and by assigned treatment group, with medical therapy divided into early and late stage (see Methods for definition of early vs. late). The mean overall survival time in the total cohort was 14.5 months ± a standard error of 0.4 months (95% confidence interval [CI] = 13.74 – 15.31 months). The mean overall survival time was shortest in the groups assigned to TTVI (13.3 ± 1.2 months, 95% CI = 11.0 – 15.6) and medical therapy with advanced stage (14.0 ± 0.5 months, 95% CI = 13.1 – 14.9). The 27 patients assigned to medical therapy with early presentation had the longest overall survival (18.1 ± 1.0 months, 95% CI = 16.1 – 20.0), followed by those assigned to surgery (15.1 ± 1,0 months, 95% CI = 13.2 – 17.0).

### TR Management in Actual Practice

Of the 470 patients retrospectively assigned to medical therapy by the ESC guidelines, 97.7% (459/470) received medical therapy, nine underwent TV surgery, and two underwent TTVI. Our center currently performs heterotopic caval valve implantation (TricValve®) on a compassionate-use basis. Of the 54 patients assigned to TV surgery, 79.6% (43/54) received medical therapy, and 20.4% (11/54) underwent TV surgery, one of which was concomitant TVR with CABG. One of the patients assigned to TTVI underwent TricValve® implantation, two underwent TV surgery, and the remaining 54 received medical therapy. In total, 556 of the patients received medical therapy, 22 underwent TV surgery, and three received TTVI during the study’s follow-up period.

## DISCUSSION

In this retrospective study of consecutive patients with more than moderate TR presenting to a tertiary care center, we showed that a minority (9.8%) could qualify for TTVI, and 76.2% had advanced left or right ventricular dysfunction or severe pulmonary hypertension precluding any surgical or transcatheter intervention. The in-hospital and 18-month mortality rates were grimly high in the advanced stage medical therapy and TTVI cohorts at 18.5%, 28.9% and 19.3%, 28.1%, respectively. Although we are unable to comment on which specific TTVI device patients are candidates for, our study sheds light on the clinical and echocardiographic characteristics of this patient population and their outcomes at a tertiary care center.

There are currently four categories of TTVI, which are at various developmental stages: coaptation devices (TriClip^®^, Pascal^®^, Dragonfly^®^, FORMA^®^), annuloplasty devices (cardioband^®^, K-Clip^®^, Trialign^®^, TriCinch^®^) orthotopic replacement (Evoque^®^, Lux^®^, Intrepid^®^) and heterotopic replacement (TricValve^®^, Trillium^®^). The first three devices aim to reduce TR severity, and the last one reduces venous congestion caused by it. There are no commercially available TR-reducing transcatheter devices in the United States, and the TricValve^®^ is only available for compassionate use. Some coaptation (TriClip^®^ and Pascal^®^) and annuloplasty devices (cardioband^®^) are commercially available in Europe, and the TriClip is expected to get approval from the Food and Drug Administration in the United States following the Trial to Evaluate Treatment with Abbott Transcatheter Clip Repair System in Patients with Moderate or Greater Tricuspid Regurgitation (TRILUMINATE) meeting its primary endpoint.^18–19^

Although several non-randomized controlled trials (RCT’s)^20–27^ using various types of TTVI devices showed symptomatic relief, the ideal timing of intervention to impact survival remains to be defined.^28–29^ The TRILUMINATE study on tricuspid edge-to-edge repair is the only RCT testing a TTVI device. ^19^ It showed that TriClip met its primary composite endpoint of all-cause mortality, tricuspid valve surgery, heart failure hospitalizations, and quality-of-life improvement using the Kansas City Cardiomyopathy Questionnaire (KCCQ) score at 12 months, but that was mainly driven by improvements in the KCCQ score. It is unclear at this stage if mortality would be reduced by Triclip after the two-year follow-up planned for this trial is completed.

While heterotopic TV replacement is a catheter-based device, it is generally a non-corrective procedure, and for the purposes of our study, we did not consider it TTVI. Except for the three patients who underwent TricValve implantation, none underwent TTVI because we did not have access to these therapies during the study period. Therefore, the 28.1% mortality rate of the patients allocated to the TTVI group reflects the untreated mortality rate of this cohort. This rate is almost threefold higher than the 10.6% mortality rate seen in the control arm of the TRILUMINATE trial, which is not surprising given the higher prevalence of comorbidities and advanced-stage disease in our TTVI group (liver cirrhosis 36.6%, CKD 65.9%, ESRD 19.3%) compared to the control group enrolled in TRILUMINATE (liver disease 9.1%, CKD 35.4%, ESRD 0%).^19^

The characteristics of the patients who are candidates for TTVI in our study mirror those of the TriValve registry patients.^30^ This registry is an ongoing, prospective database of all patients undergoing TTVI outside of RCT’s. In 2017, they reported on 107 patients with clinically significant TR treated with various TTVI devices, mainly the MitraClip device. The mean age was 76 years, atrial fibrillation 79%, female 60%, ascites 27%, NYHA class III or IV 95%, prior left-sided valve surgery 26%, mean LVEF 51%, and mean TAPSE 16 mm. These demographics and clinical variables were very similar to our TTVI patient cohort; mean age of 75 years, atrial arrhythmia 79%, female 56%, ascites 35%, NYHA class III or IV 82%, prior valve surgery 26%, mean LVEF 54%, and mean TAPSE 20 mm.

The TTVI treatment group in our study was derived from a pool of patients who presented to our tertiary care center with an already high burden of comorbidities and end-organ disease (liver cirrhosis 17.7%, CKD 64.0%, ESRD 19.8%) and a 26.7% mortality rate. There is a direct association between TR severity and mortality^8, 31–33^, but comorbidities, which are indicators of late presentation and end-organ failure, predict mortality independent of TR severity.^17^ Breitninger et al. showed that age, male gender, renal dysfunction, presence of CHF, liver disease, lung disease, and TR severity were independently associated with mortality.^22^ Topilsky et al. showed that left sided valvular disease and LV systolic dysfunction were associated with the poorest survival in patients with functional TR, followed by pulmonary hypertension and primary TR.^17^

As discussed above, our study’s patient population had a high burden of comorbidities and signs of end-stage disease than the TRILUMINATE population. Therefore, TTVI performed at the advanced stage noted in our inpatient cohort may not impact survival although it could improve quality of life. A suggestion that this might be the case comes from the Mayo Clinic study by Breitinger et al. done in 13,608 with moderate to severe TR and a median follow-up of 6.5 years.^33^ In patients with significant comorbidities, the severity of TR was not associated with differences in survival. In contrast, in those with fewer comorbidities, increasing TR severity was associated with worse survival. Another study showed similar results. Neuhold et al. studied 576 patients with heart failure and noted that TR severity was predictive of poor outcomes in those with mildly to moderately impaired LV function but not in those with a severely depressed EF.^34^ Nonetheless, it is reassuring that the TriValve registry reported a low all-cause 30-day mortality rate of 3.7% in 106 patients who underwent TTVI, despite a high prevalence of comorbidities.^30^ It is unclear, however, if there are unreported factors that led to a selection bias in the TriValve registry, resulting in a more favorable 30-day mortality rate.

Tricuspid regurgitation is a complex disease with etiology having an impact on management and outcomes. Most TR is functional, thus the rate of progression and success of treatment depends on non-tricuspid valve drivers, such as aortic and mitral valvular disease, ventricular function, and arrhythmia control.^35^ Clinical trials of TTVI devices aiming to reduce TR will need to account for these diseases. Adding to the complexity is the fragile nature of the right ventricle that is not designed to withstand chronic pressure and volume overload. Seventy six percent of the patients in our study had disease too advanced for any intervention with a mortality rate just under 30% by 18 months. This rate stresses the importance of early detection that should initiate a comprehensive management plan for all factors contributing to TR, and early referral for intervention.^36^

Therefore, future clinical trials using TTVI devices should not only focus on patients with advanced stages but also on those at earlier or even asymptomatic stages of TR where early intervention may confer a survival benefit in addition to symptomatic benefit.^37^ The Mayo Clinic reported that patients with a low TRIO (Tricuspid Regurgitation Impact on Outcomes) score with few comorbidities might benefit the most from interventions to correct TR, based on 10-year all-cause mortality.^33^ Provided that evidence of improved survival may be realized with longer follow-up, we expect that TTVI guidelines would eventually evolve to include lower-risk patients, analogous to the evolution of the guidelines for the transcatheter aortic valve implantation (TAVI) procedure.

Limitations of this retrospective, single-center analysis merit consideration. NYHA functional class was not defined for over half of our cohort, and thus physical exam history of lower extremity edema or ascites was used to indicate signs of RHF instead. Due to differences in patients’ transthoracic echocardiogram imaging, some echocardiographic parameters, such as the S’ or TAPSE were not measurable for all patients. Furthermore, the ESC 2021 guidelines do not specifically define parameters for determining appropriateness for surgery, so we used our own criteria, which may have underestimated the number of patients who were too high risk for surgery.

The present study findings show that most patients presenting to a tertiary care center with more than moderate TR are too advanced for any intervention and have high in-hospital and late mortality rates. We illustrated that a minority of these patients would clinically qualify for TTVI. There is a need to improve care for patients with TR through earlier detection that should initiate a comprehensive management plan for all factors contributing to TR, and early referral for intervention. Future clinical trials with longer follow up will be needed to assess TTVI devices in patients at earlier or even asymptomatic stages of TR when there may be a greater likelihood of improving survival.

## Data Availability

The data that support the findings of this study are available on request from the corresponding author, F.A.M

## ACKNOWLEDGEMENTS

The authors acknowledge the invaluable assistance of the structural cardiology team at the Tampa General Hospital Heart and Vascular Institute in the conduct of this study. Conceptualization and methodology: A.O.D.C., C.B.S, D.R.C., and F.A.M. Investigation: A.O.D.C., C.B.S, S.N.M., and J.A.M. Formal analysis and interpretation: A.O.D.C., C.B.S, D.R.C., and F.A.M. Writing: A.O.D.C., C.B.S, and F.A.M. Writing–review and editing and supervision: A.O.D.C., C.B.S, S.N.M., J.A.M., D.R.C., and F.A.M.

